# Thanzi La Mawa (TLM) datasets: health worker time and motion, patient exit interview and follow-up, and health facility resources, perceptions and quality in Malawi

**DOI:** 10.1101/2024.11.14.24317330

**Authors:** Dominic Nkhoma, Precious Chitsulo, Watipaso Mulwafu, Emmanuel Mnjowe, Wiktoria Tafesse, Sakshi Mohan, Timothy B Hallet, Joseph H Collins, Paul Revill, Martin Chalkley, Victor Mwapasa, Joseph Mfutso-Bengo, Tim Colbourn

## Abstract

The Thanzi La Mawa (TLM) study aims to enhance understanding of healthcare delivery and resource allocation in Malawi by capturing real-world data across a range of health facilities. To inform the Thanzi La Onse (TLO) model, which is the first comprehensive health system model developed for any country, this study uses a cross-sectional, mixed-methods approach to collect data on healthcare worker productivity, patient experiences, facility resources, and care quality. The TLM dataset includes information from 29 health facilities sampled across Malawi, covering facility audits, patient exit interviews, follow-ups, time and motion studies, and healthcare worker interviews, conducted from January to May 2024.

Through these data collection tools, the TLM study gathers insights into critical areas such as time allocation of health workers, healthcare resource availability, patient satisfaction, and overall service quality. This data is crucial for enhancing the TLO model’s capacity to answer complex policy questions related to health resource allocation in Malawi. The study also offers a structured framework that other countries in East, Central, and Southern Africa can adopt to improve their healthcare systems.

By documenting methods and protocols, this paper provides valuable guidance for researchers and policymakers interested in healthcare system evaluation and improvement. Given the formal adoption of the TLO model in Malawi, the TLM dataset serves as a foundation for ongoing analyses into quality of care, healthcare workforce efficiency, and patient outcomes. This study seeks to support informed decision-making and future implementation of comprehensive healthcare system models in similar settings.

## A. Introduction

A major challenge in health systems of Low- and Middle-Income Countries such as Malawi is understanding the impact of the investments being made across the health system and disease areas, and using this, being able to model future impacts of alternative resource use and design options. Through <u>Thanzi La Onse (TLO, 2018 - 2021)</u> and <u>Thanzi La Mawa (TLM, 2021-2025)</u> a detailed individual-based mathematical model of health, disease and healthcare costs across the life-course has been developed to answer resource allocation policy questions whilst considering the whole health system in Malawi.^1^ The TLO model is the first all-diseases-whole health-system model for a country ever developed. The model is accessible via www.tlomodel.org. The first phase of the model development focused on characterising the Malawi health system with available survey and administrative datasets. Where no data could be found for Malawi, proxies were used. Datasets like DHIS2^2^, Harmonized Health Facility Assessment (HHFA)^3^, Demographic and Health Surveys (DHS)^4^, Service Provision Assessments (SPA)^5^ and Service Availability and Readiness Assessment (SARA)^6^ /Service Delivery Indicator (SDI)^7^ surveys have been used and have provided valuable insights. The SPA provides structured information on a small number of patient consultations (family planning, under five care, antenatal care (ANC) and delivery). However, overall these pre-existing datasets provide inadequate granularity and lack detail necessary to represent service delivery, health worker productivity, and patient interactions, for example, in the TLO model.. The benefit of the surveys and datasets described in this paper are that they capture information on; 1) all activities (minute-by-minute) carried out by a large number of different types of cadres and clinics (cadres and clinics not included in other surveys) during the course of multiple days/shifts; 2) the components of care provided across different patients and health conditions, including patient exit and follow-up at two weeks; and 3) daily patient volumes by clinics/departments including healthcare worker supply by clinics/departments (allowing capture of heterogeneity by clinic/ward within facilities).

This paper provides an overview of the TLM survey, data collection methods, data collection procedures, data modules and core variables, and quality assurance measures and expected analytical outputs from the survey. Documenting this provides researchers with an overview of the data and provides detailed background for forthcoming analyses of healthcare worker time and motion, task shifting, quality of care and related work. Given the formal adoption of the TLO model in Malawi and the growing interest across African countries, particularly within the East, Central, and Southern Africa (ECSA) region for also adopting its approach, documenting this survey protocol is also crucial for guiding future implementations and expanding the model’s use in healthcare system improvement. The TLM survey has the unique advantage of describing the same processes from multiple angles: the facilities and staff available, the demand on their times, the needs of patients, how time is used in practice, and patient experiences.

## B. Materials and Methods

The survey used a cross-sectional, mixed-methods approach to collect data in 29 health facilities across Malawi between January and May 2024. Specifically, the survey included facility audits, time-motion, patient exit and follow-up, and qualitative interviews with facility managers to capture a detailed picture of service delivery and resource use across primary facilities (health centres and community hospitals), secondary hospitals, and tertiary hospitals following the official categorization of the Malawi Ministry of Health (MOH 2023)^8^ adopted in the TLO model.

Health services in Malawi are delivered at 1) the community level and health posts (the community health system), 2) Health Centres and Community Hospitals (primary facilities), 3) Secondary Hospitals, and 4) Tertiary (MOH 2023)^8^. The 2023

Master Health Facility Register^9^ was used as the primary sampling frame and facility details were triangulated with the Health Management Information System (MOH DHIS2)^10^. There were 1,424 facilities on the final list; of these, 2 facilities did not have enough information to facilitate identification of facility type. Table 1 shows the distribution of the 1,422 facilities classified by the above levels of healthcare delivery system. For this survey, we focused on Government and CHAM/IHAM health facilities because that is the primary focus of the TLO model. Table 2 shows the final sampling framework by facility ownership (Government and CHAM/IHAM), and location (rural/urban) – please note a few facilities did not have information on rural/urban in the master list hence they are not in Table 2.

**Table 1:** Distribution of health facilities in Malawi by facility ownership.

| Facility Type | Facility Ownership |  |  |  | Grand Total |
| --- | --- | --- | --- | --- | --- |
|  | Government | CHAM/IHAM | NGO | Private |  |
| Central Hospital | 5 |  |  |  | 5 |
| Secondary Hospital | 24 | 19 | 4 | 17 | 64 |
| Rural/Community Hospital | 20 | 35 |  |  | 55 |
| Health Centre | 382 | 124 | 17 | 103 | 626 |
| Health Post | 250 | 29 | 58 | 335 | 672 |
| **Grand Total** | **681** | **207** | **79** | **455** | **1422** |

**Table 2:** final health facility sampling framework.

| Facility Type | Government |  |  | CHAM/IHAM |  |  | Grand Total |
| --- | --- | --- | --- | --- | --- | --- | --- |
|  | Rural | Urban | Total | Rural | Urban | Total |  |
| Central Hospital |  | 5 | 5 |  |  |  | 5 |
| Secondary Hospital | 3 | 21 | 24 | 11 | 8 | 19 | 43 |
| Rural/Community Hospital | 18 | 2 | 20 | 32 | 2 | 34 | 54 |
| Health Centre | 355 | 26 | 381 | 102 | 18 | 120 | 501 |

<sup>9</sup> Ministry of Health Malawi. (n.d.). *Zipatala health facilities*. Retrieved November 13, 2024, from https://zipatala.health.gov.mw/facilities
<sup>10</sup> Ministry of Health, Malawi. (n.d.). *DHIS2 Health Information System*. Retrieved November 13, 2024, from https://dhis2.health.gov.mw

|  |  |  |  |  |  |  |  |
| --- | --- | --- | --- | --- | --- | --- | --- |
| **Grand Total** | **376** | **54** | **430** | **145** | **28** | **173** | **603** |

A total of 174 health workers and 3,480 patients were selected from 29 sampled facilities. To ensure a representative sample, facilities were stratified first by ownership category (Ministry of Health [MOH] and Christian Health Association of Malawi [CHAM]). Within each ownership category, facilities were further stratified by urban and rural location. Each location category was then stratified by patient volume, with facilities randomly selected via a random number generator in Microsoft Excel from each of the stratified lists. Using this stratification approach, out of the 29 facilities, 24 (primary and secondary facilities) were randomly selected, including 12 urban and 12 rural facilities, evenly divided between MOH and CHAM (6 each). For each ownership category, the 6 sampled facilities consisted of 2 health centers, 2 rural hospitals, and 2 secondary hospitals. For each facility type, one high-volume and one low-volume facility were selected. All 5 central hospitals were included in the survey to capture insights across facility levels. Each facility was surveyed continuously for a period of 10 to 28 days, with 2 to 7 nights included in the data collection, depending on facility type (as detailed in Table 3).

**Table 3:** Number of days and nights at each facility type.

| Facility type | Total number of days there | Consecutive days of time and motion data collection | Day off to switch from day to night | Consecutive nights of time and motion data collection |
| --- | --- | --- | --- | --- |
| Health centre | 10 | 7 | 1 | 2 (if health centre opens at night including on-call) |
| Community Hospital | 12 | 9 | 1 | 2 |
| District hospital | 14 | 10 | 1 | 3 |
| Central hospital | 28 | 10, then 1 day off, then another 9 | 1 | 7 |

Patients were randomly selected based on their time of attendance, and health workers were observed across different cadres. Our proposed sample size was arrived at to have sufficient precision to inform parameters of our mathematical model. Our target sample size of 174 health workers and 3480 patients (in 29 facilities) was calculated to allow us characterise demand and resource use for informing the model analyses (no such data are available currently). For example, one key parameter is the proportion of patients that attend who are 16–49-year-old men, and the proportion of those that are treated for HIV. If 400 randomly-sampled patients are 16–49-year-old men and 40 of those are being treated for HIV, then these parameters are estimated to be 16.7% (95%CI: 15.2%, 18.2%) and 10% (95%CI: 7.2%, 13.4%), respectively. Another key parameter is the proportion of patients that nurses at district hospitals see who have diabetes. If 32 nurses are followed for a day during the time-and-motion study at district hospitals and see a total of 512 patients of which 128 are diabetes patients, then it can be estimated that 25% (95%CI: 21.3%, 29.0%) of the patients that nurses see per day have diabetes.

## C. Survey Instruments Used

The TLM facility survey utilized six inter-locking complementary data collection tools to capture comprehensive insights into healthcare service delivery.

Figure 1 demonstrates this, illustrating how these different tools help characterize the same process from different angles. <u>Tool 1</u>, the Health Facility Audit, assessed the availability of critical resources, including human resources, equipment, and drugs. <u>Tool 2</u>, the Patient Exit Survey, gathered information on patient demographics, healthcare-seeking behaviour, and satisfaction with the services obtained. <u>Tool 3</u>, the Time and Motion Study, recorded minute-by-minute activities of health workers to measure time use. <u>Tool 4</u>, the Qualitative Interviews, explored healthcare workers’ perspectives on resource challenges, service delivery, and motivation. <u>Tool 5</u>, was used to follow-up on patients two weeks after patient exit interviews to measure self-assessed health outcomes and subsequent healthcare use after their facility visit. <u>Tool 6</u>, was used to record opening and closing times of clinics in facilities as well as number of healthcare workers and patients seen.

**Figure 1:**
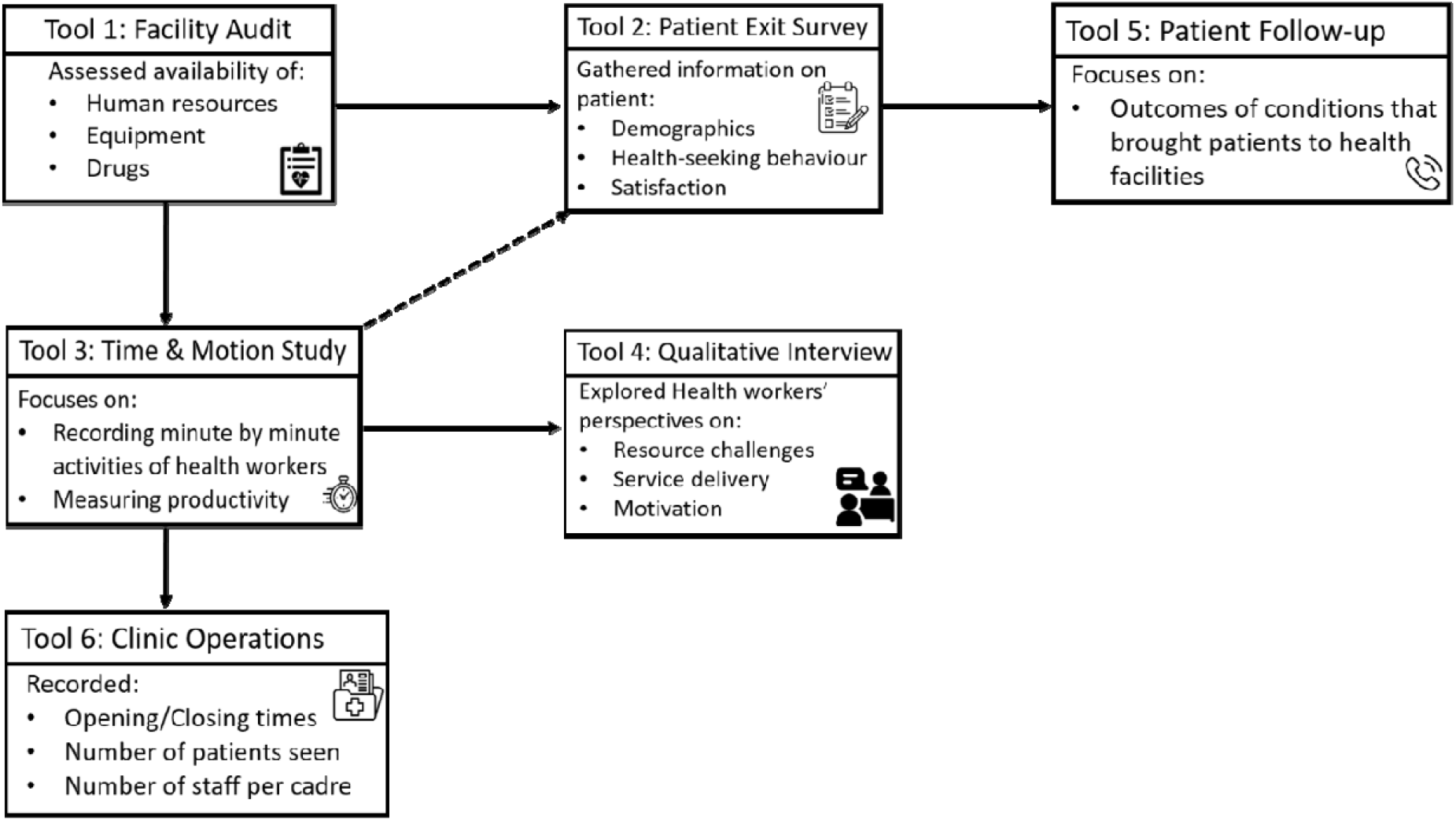
TLM data collection tools

### Pretesting and Training

Before full-scale data collection, a comprehensive training program was conducted from January 15 to January 22, 2024. The training involved piloting data collection tools first at Kabudula Community Hospital, Lilongwe district followed by debriefing sessions to incorporate feedback and make necessary adjustments, then piloting at Area 25 Health Centre in Lilongwe followed by another debrief session. Enumerators were trained in both quantitative and qualitative data collection methods, including the use of KoboToolbox, an online platform for real-time data entry. Emphasis was placed on ensuring that data collectors understood the tools, ethical considerations, and strategies to minimize bias.

### Field Teams and Management

Data collection was carried out by five teams, each consisting of three enumerators each with at least a Bachelor’s Degree, supervised by a lead supervisor. Each team had 2 medically trained enumerators that carried out time and motion observations and one enumerator not medically trained. Field management included regular reporting and troubleshooting through WhatsApp groups and daily progress tracking on Google Sheets. Supervisors conducted periodic on-site visits to monitor data quality and provide real-time feedback to teams. This system ensured timely identification and resolution of issues during data collection.

### Data Management

Data was entered into tablets using KoboToolbox and uploaded to a secure, password-protected server managed by the Kamuzu University of Health Sciences (KUHES), Health Economics and Policy Unit (HEPU). Each entry was cross-checked by supervisors before final approval. Data cleaning involved checking for outliers and addressing any inconsistencies identified during initial analysis.

## 6. Data (and Software) Availability

The datasets can be accessed from a GitHub repository through the link (https://github.com/HEPUMW/TLM-Study-Data). From each of the data collection tools there is a dataset or two as follows:

### A. Tool 1 - Facility Audit and Human Resources for Health (HRH)

This dataset provides detailed information on healthcare facilities, capturing a wide array of health services, facility management, resource availability, and financial data. It includes fields such as facility type, district, and a catalogue of clinic-specific services that are available (e.g., HIV clinic, ANC clinic, TB clinic), with the days these services are operational. Other aspects recorded include the availability of surgical theatres and beds for different patient types (e.g., paediatric, neonatal, emergency). Additionally, the dataset captures financial information about the facility, including sources of funds, costs related to facility operations such as electricity, water, cleaning, and medical supplies. There are also sections related to the availability of equipment and stockouts of medicines and other consumables. This detailed dataset can be used to evaluate the efficiency of healthcare services, identify gaps in resource distribution, and support decisions to improve the management of health facilities.

The dataset includes information on the workforce for each health facility. For each of the 27 categories of cadres included, it captures the estimated number of full-time, part-time, and temporary positions that are funded in principle alongside whether these positions are filled. Additionally, it provides information on duty and on-call schedules across weekdays, weekends, and nights, with data on whether staff were assigned duties during these periods. This information is critical for evaluating the staffing structure, workload distribution, and operational capacity of the healthcare facility. It also shows how different cadres contribute to coverage at various times, shedding light on the overall workforce availability at the facility.

This information will help inform parameters on the types of appointments within the health system, the availability of consumables, the number of beds available in each ward well as whether or not each appointment is offered at each facility level in the Thanzi la Onse model.

### B. Tool 2 – Patient Exit Survey

The dataset captures detailed patient exit survey information from sampled health facilities, focusing on demographics, service utilization, and patient satisfaction. Patients were surveyed at random as they left the facility (data collectors were located at the facility gates and asked exiting patients if they would like to complete the exit survey). The patient exit survey includes information on the age, gender, reason for visiting the facility, and whether the patient received care. It also records the time of arrival and departure, the clinic they attended, and any specific medical conditions they may have had. In addition, the dataset tracks patient feedback on their healthcare experience, including perceived respect from staff, consultation quality, waiting times, and treatment availability. Costs (direct monetary costs and associated opportunity costs) incurred by the patients are also recorded, including transportation costs, food, and accommodation, alongside details about household assets. This dataset offers rich insights into patient experiences and satisfaction levels, capturing the socio-economic context and the quality of care provided at the health facility. The variety of information recorded highlights the facility’s operational aspects and provides a valuable foundation for understanding and improving healthcare service delivery in the area.

### C. Tool 3 – Activity Level Time Motion Study

The dataset represents an activity-level Time and Motion Survey (TMS) conducted in sampled health facilities. It focuses on tracking activities of healthcare workers across a wide range of clinics/departments, with details captured on the exact time (to the nearest minute) spent on various tasks, patient interactions, and healthcare provision. Each row in the dataset tracks a specific instant of time in minutes, noting variables such as the health condition addressed, the specific activity (there were around 37 activity options to choose from for every clinic in the tool), and whether the patient is new or a returning patient. Unique patient ID numbers were handed out after a patient exited each observed (timed) consultation in order to link the observed service provision to patient outcomes at follow-up (i.e. to link the patient to the patient exit survey - Tool 2 - conducted as they left the facility)

This dataset quantifies health care worker time use by clinic, health condition treated and activity done, by healthcare worker cadre. It will be used for analyses of health worker time use and productivity, and of task-shifting: different health worker cadres covering activities originally intended for other cadres.

This information informs parameters on the time taken for each appointment, according to officer and facility type in the model.

### D. Tool 4 – Healthcare Worker Interview

This covers the qualitative interviews with health care workers. These semi-structured interviews covered functioning of clinics, barriers to high quality health care provision, use of cash budgets, leakages, prioritisation of patients, and motivation. The purpose of these interviews is to explore and investigate things we don’t know, as well as to provide explanation of things we think we know, and to validate findings from our quantitative data.

h

### E. Tool 5 – Patient Follow Up

Tool 5 was administered as a phone call interview to follow up all patients two weeks after being interviewed using Tool 2. After the interview, the enumerators requested the survey respondent’s phone number; if they didn’t have one, then they provided the number for next of kin or neighbours. The follow-up survey tracks key outcomes related to patient recovery after receiving treatment, specifically focusing on whether the patient’s illness has resolved, their current health status, and whether the treatment administered was effective. A total of 2,875 individuals (out of 4181 patients surveyed) had their contact details collected for the follow-up survey out of which, 2,711 follow-up interviews were successfully conducted (65% of the 4181 patients originally surveyed - Tool 2).

### F. Tool 6 – Clinic Operations

This dataset records information on opening hours, number and type of healthcare workers and number of patients seen verified by records by clinic in each facility. Important parameters such as the daily capabilities in terms of minutes of time available of each type of officer in each facility will be informed by this information in the model among others.

## 7. Reporting Guidelines

Not applicable as we are not reporting study results in this paper.

## 8. Consent

Written informed consent for involvement of patients in facility exit as well as follow up interviews, health workers in observational time and motion study as well as facility in-charges in the facility audit interviews was obtained from the participants. Where it could not be obtained directly from the participants, especially in the case of young patients, guardians provided the consent. All participants were informed of the voluntary nature of their involvement and were assured their information would be kept strictly confidential and anonymous.

## Data Availability

This paper describes the surveys done to create the data. We anticipate our data repository will be publicly available in May 2025 after we have completed analysis of the data.

## 9. Author Contributions

Conceptualization: DN, PC, WT, VM, JMB, TC

Data Curation: PC

Formal Analysis: no analysis in this paper, which describes datasets Funding Acquisition: TBH, PR, MC, JMB, TC

Investigation: DN, PC, WM, EM, WT, VM, JMB, TC

Methodology: DN, PC, WT, SM, VM, JMB, TC

Project Administration: DN, VM, JMB

Resources: PC, WM, EM, TC

Software: PC

Supervision: DN, PC, WM, EM, VM, JMB

Validation: DN, PC, WM, EM, WT, SM, TBH, JC, PR, MC, VM, JMB, TC

Visualization: DN, PC

Writing – Original Draft Preparation: DN, PC, TC

Writing – Review & Editing: DN, PC, WM, EM, WT, SM, TBH, JC, PR, MC, VM, JMB, TC

## 12. Acknowledgements

We would like to extend our heartfelt gratitude to the Health Economics and Policy Unit (HEPU) team for their invaluable support in the preparation of this paper. The contributions of HEPU members through their constructive feedback, insightful comments, and expert guidance have greatly enhanced the quality and coherence of our work. We are especially thankful for HEPU’s instrumental role in organizing the Thanzi La Mawa (TLM) think-tank meeting, which brought together key stakeholders from the health sector. This forum provided an essential platform to discuss the Thanzi La Onse (TLO) model and explore its potential to improve health outcomes. The presentation of the TLM study overview at this meeting formed the foundation of this paper and offered critical insights that informed our analysis and approach.

We deeply appreciate the collective effort and dedication of all HEPU employees, whose contributions have been invaluable in shaping this research.

## Footnotes

1 Hallett TB, Mangal TD, Tamuri AU, et al. Estimates of resource use in the public-sector health-care system and the effect of strengthening health-care services in Malawi during 2015–19: a modelling study (Thanzi La Onse). *The Lancet Global Health* 2024.

2 Ministry of Health, Malawi. (n.d.). *DHIS2 Health Information System*. Retrieved November 13, 2024, from https://dhis2.health.gov.mw

3 Malawi - *Harmonized Health Facility Assessment*: 2018-2019 Report : Main Report (English). Washington, D.C. : World Bank Group. http://documents.worldbank.org/curated/en/417871611550272923/Main-Report

4 The DHS Program. (n.d.). *Malawi: Standard DHS, 2022*. Retrieved November 13, 2024, from https://dhsprogram.com/Countries/Country-Main.cfm?ctry_id=24&c=Malawi&Country=Malawi&cn=&r=1

5 The DHS Program. (n.d.). *Malawi: Standard DHS, 2022*. Retrieved November 13, 2024, from https://dhsprogram.com/Countries/Country-Main.cfm?ctry_id=24&c=Malawi&Country=Malawi&cn=&r=1

6 World Health Organization. (n.d.). *Service Availability and Readiness Assessment (SARA)*. Retrieved November 13, 2024, from https://www.who.int/data/data-collection-tools/service-availability-and-readiness-assessment-(sara)

7 World Bank. (n.d.). *Service Delivery Indicators: Health - Country reports and data*. Retrieved November 13, 2024, from https://www.worldbank.org/en/programs/service-delivery-indicators/health/country-reports-and-data

8 Ministry of Health Malawi. (2023). Health sector strategic plan (HSSP) III. Ministry of Health Malawi

9 Ministry of Health Malawi. (n.d.). *Zipatala health facilities*. Retrieved November 13, 2024, from https://zipatala.health.gov.mw/facilities

10 Ministry of Health, Malawi. (n.d.). *DHIS2 Health Information System*. Retrieved November 13, 2024, from https://dhis2.health.gov.mw

